## Supplemental Material for "AI-DRIVEN QUANTIFICATION OF GROUND GLASS OPACITIES IN LUNGS OF COVID-19 PATIENTS USING 3D COMPUTED TOMOGRAPHY IMAGING"

**Monjoy Saha[1], Sagar B. Amin[2], and Ashish Sharma[1]**
[1] Department of Biomedical Informatics
[2] Department of Radiology and Imaging Sciences
Emory University School of Medicine, Atlanta, GA 30322, USA
&
**T. K. Satish Kumar[3] and Rajiv K. Kalia[3, 4, *]**
[3] Department of Computer Science
[4] Collaboratory for Advanced Computing and Simulations
[4] Department of Physics & Astronomy
[4] Department of Chemical Engineering and Materials Science
[4] Department of Biomedical Engineering
University of Southern California, Los Angeles, CA 90089, USA

### SUPPLEMENTARY MATERIAL

In this section, we describe concepts such as thresholding, dilation, erosion, and structuring element operation which have been used to segment lungs and GGOs. The supplementary material also includes the convex hull algorithm for GGO segmentation and the Cox model for automated classification of abnormalities seen in CT lung scans of COVID-19 patients. Additional PointNet++ results for GGOs and movies of GGO segmentation are also presented in this section.

#### CONVEX HULL ALGORITHM

Over the years, several algorithms have been developed to efficiently find the convex hull of a set of coplanar points. For a set of $N$ unstructured points, earlier developers of convex hull algorithms were able to reduce the complexity to O($N$ log$N$). If the points are structured, e. g., the vertices of a non-self-intersecting polygon given in a clockwise or anti-clockwise sequence, it's possible to reduce the complexity of the algorithm to O($N$) [2, 5].

We use a global convex hull (GCH) algorithm to find the convex hull of unstructured coplanar points. The polygon constructed by the algorithm consists of a finite number of linear segments forming a Jordan curve. The GCH algorithm is an extension of a simpler algorithm which has four steps:

1. Find four extremal vertices on the left ($V_L$), right ($V_R$), top ($V_T$) and bottom ($V_B$). Take one of them, say $V_L$, and suppose $V_1 = V'_k$. Then let $V_2 = V'_{k+1}, \dots\dots, V_m = V'_{m+k-1\ mod\ m}$.
2. Compute the following quantity,

$$S_i \cong (x_{i+1} - x_{i-1})(y_{i-1} - y_i) - (y_{i+1} - y_{i-1})(y_{i-1} - y_i)$$

   where ($x_i$, $y_i$) are the Cartesian coordinates of $V_i$. If $S_i < 0$, $V_i$ is a vertex and keep it. If $S_i >$ 0, remove it and find another nonconvex vertex.

- 

3. Stop removing nonconvex vertices when $V_1$ is reached again because $V_1$ is an extremal vertex.
4. Repeat steps 1, 2, and 3 for the remaining three extremal vertices.

This algorithm can sometimes lead to non-simple or self-intersecting polygons The GCH algorithm was designed to address this problem and can produce a non-self-intersecting polygon as a convex hull.

#### Cox Proportional-Hazards Model

Cox model is a widely used statistical technique to determine the effect of many explanatory variables on the survival of a patient. Like all survival models, it consists of a baseline hazard function and another function that describes the effect of hazard parameters in terms of explanatory covariates of a patient. The baseline function, $\lambda_0(t)$, describes changes in the risk of an event over time at the baseline level of covariates of a patient $i$: $X_i = \{ X_{i1}, X_{i2}, \ldots. X_{in}\}$. The covariates may include a patient's age, gender, medical preconditions and other diseases at the start of study as well as the treatment. The Cox model is based on the condition that covariates are independent and multiplicatively related to the hazard. For a patient with covariates $X_i$, the form of the hazard function at time $t$ is

$$\lambda(t|X_i) = \lambda_0(t)exp\left(\sum_j \beta_j X_{ji}\right)$$

The probability of an event occurring at time $\tau$ is given by,

$$p_i = \frac{\lambda(\tau_i|X_i)}{\sum_{j,\tau_j>\tau_i}\lambda(\tau_i|X_j)} = \frac{q_i}{\sum_{j,\tau_j>\tau_i} q_j}$$

where $q_i = exp\left(\sum_j \beta_j X_{ji}\right)$. Taking into account all the patients treated independently, the probability of occurrence of an event is given by $P(\beta) = \prod_\beta p_i$, where $\beta$ stands collectively for all the variables $\beta_i$. In the model, the log-likelihood of the probability function

$$L(\beta) = \sum_i X_i\beta - log \sum_{j,\tau_j>\tau_i} q_j$$

is maximized with respect to all the $\beta_i$ variables.

#### Intersection over Union (*IoU*)

*IoU*, also known as the Jaccard index, is commonly used in segmentation, object detection and tracking tasks. Object detection involves finding the location of an object by drawing a box around it and assigning a class to that object. *IoU* for two objects $A$ and $B$ is computed from the ratio of the intersection, $I$, and union, $U$, between $A$ and $B$:

$$IoU =. \frac{|I|}{|U|} = \frac{|A\cap B|}{|A\cup B|}$$

#### Thresholding

A threshold is a cut-off value, which has two regions, i.e., above and below the cut-off value. In image processing, thresholding is used to segment the region of interest.

#### Dilation and Erosion

The two most significant morphological operations are dilation and erosion. Dilation and erosion are used to perform two different tasks. Dilation adds pixels in an image, and erosion removes pixels from an image.

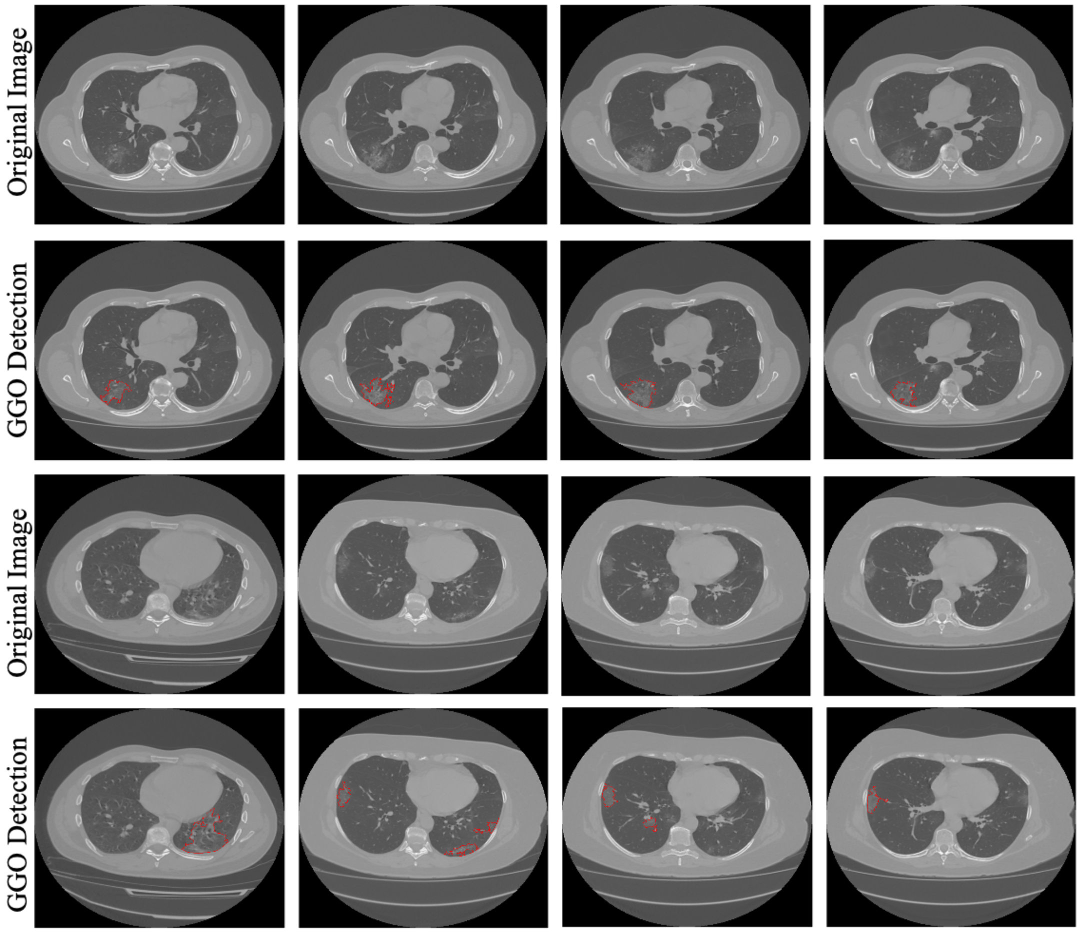


**Figure 1S:** GGO detection results. Red color regions indicate GGOs.

#### STRUCTURING ELEMENT OPERATION

The structuring element is a shape matrix, which is used for processing pixels in the region of interest.

#### PIXEL INTENSITIES TO HOUNSFIELD UNITS (HU) CONVERSION

The conversion is done with the following equation: $HU(y) = slope\ .y + intercept$, where $y$ denotes the pixel value.

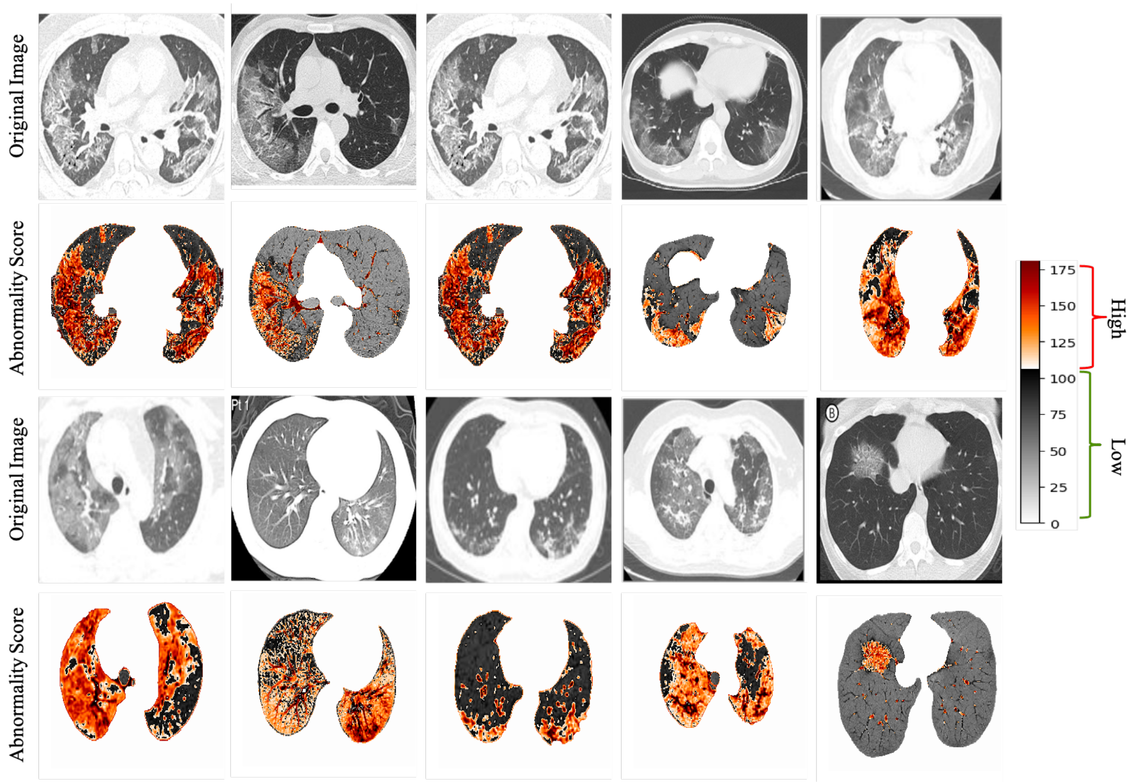


**Figure 2S:** Shows predicted high- and low-abnormality regions computed by the CNN network and Cox proportional hazard model. The range of colors is shown in the color color bar. The abnormality score of small regions of lung images has been computed and represented using a heatmap. The malignant and GGO regions have been identified as high-abnormality regions. Similarly, healthy and non-malignant regions have been identified as low-abnormality regions. This approach doesn't give details of grade or any other pathological findings. A portion of an infected lung may have healthy or non-malignant regions and vice versa. We are just identifying those abnormalities in lung images.

### POINTNET++ RESULTS - VIDEO LINK

https://drive.google.com/drive/folders/1q8U5JesS6DgStYXohaGisl2y39rSS4RC?usp=sharing

**References of Supplementary Material**:


1. Kennedy, A., Dowling, J., Greer, P.B. and Ebert, M.A., 2018. Estimation of Hounsfield unit conversion parameters for pelvic CT images. Australasian physical & engineering sciences in medicine, 41(3), pp.739-745.
2. Spruance, S.L., Reid, J.E., Grace, M. and Samore, M., 2004. Hazard ratio in clinical trials. Antimicrobial agents and chemotherapy, 48(8), pp.2787-2792.
